# Correlation Between Blood and CSF Compartment Cytokines and Chemokines in Subjects with Cryptococcal Meningitis

**DOI:** 10.1101/2020.05.08.20095232

**Authors:** Elizabeth C Okafor, Katherine H Hullsiek, Darlisha A Williams, James E Scriven, Joshua Rhein, Henry W Nabeta, Abdu K Musubire, Radha Rajasingham, Conrad Muzoora, Charlotte Schutz, Graeme Meintjes, David B Meya, David R Boulware

## Abstract

**Background:** Though peripheral blood is a crucial sample to study immunology, it is unclear whether the immune environment in the peripheral vasculature correlates with that at the endorgan site of infection. Utilizing cryptococcal meningitis as a model, we investigated the correlation between serum and cerebrospinal fluid biomarkers over time.

**Methods:** We analyzed the cerebrospinal fluid and serum of 160 subjects presenting with first episode cryptococcal meningitis for soluble cytokines and chemokines measured by Luminex assay. Specimens were collected at meningitis diagnosis, 1-week, and 2-weeks post cryptococcal diagnosis. We compared paired samples by Spearman correlation and the p-value was set at <0.01.

**Results:** Of the 21 analytes tested at baseline, there was no correlation detected between nearly all analytes. A weak negative correlation was found between serum and cerebrospinal fluid levels of interferon-gamma (Rho= −0.214, p= .007) and interleukin-4 (Rho= −0.232, p=. 003). There was no correlation at 1-week post cryptococcal diagnosis. However, at 2-weeks post diagnosis, there was a weak positive correlation between levels of granulocyte-macrophage colony-stimulating factor (Rho= 0.25, p= .007) between serum and cerebrospinal fluid. No cytokine or chemokine showed consistent correlation overtime.

**Conclusion:** Based on our analysis of 21 biomarkers, serum and cerebrospinal fluid immune responses do not correlate. There appears to be a distinct immune environment in terms of soluble biomarkers in the vasculature versus end-organ site of infection. While this is a model of HIV-related cryptococcal meningitis, we postulate that assuming the blood compartment is representative of the immune function at the end-organ site of infection may not be appropriate.

## INTRODUCTION

Immunologic investigations are critical to understanding the pathogenesis of infectious disease. Often human or animal subjects have their blood drawn to assess cell phenotype, function, or soluble cytokines or chemokines in order to characterize the immune response to infection. Blood is a convenient and available compartment to analyze. However, it is unclear if analysis of cells trafficking in the peripheral vasculature reflects the actual immune response at the end organ site compartment (e.g. CSF in meningitis, lung in pneumonia). This is important because the end organ site of infection is often the primary site from which initial or disease-specific signs and symptoms manifest. Of even more concern is the possibility of unknowingly overestimating or underestimating the immune response based on analysis of immune cells in the vasculature.

Analysis of site-specific immune responses in humans is difficult but not impossible. Cryptococcal meningitis is one model which allows investigation of the compartment-specific immune responses. In cryptococcal meningitis, frequent therapeutic lumbar punctures to control intracranial pressure provide a large volume of CSF over the course of treatment. Analysis of CSF provides information about the immune responses in the central nervous system (CNS). Yet, cryptococcal meningitis is not solely a localized CNS infection, but a systemic infection whereby inhaled cryptococcal spores germinate in the lung and yeast cells disseminate via the blood throughout the body [1, 2]. Peripheral blood mononuclear cells, serum, and plasma provide information about immune responses in the peripheral vascular compartment.

In cryptococcosis, but more broadly in infectious disease, the correlation between the immune response in the peripheral vasculature versus immune response in the end-organ compartment is poorly characterized. The objective of the present analysis is to determine the correlation between 21 peripheral blood and CSF cytokines and chemokines in a cohort of subjects with first episode cryptococcosis. In addition, we report the correlation results over time using three distinct timepoints during the course of cryptococcosis treatment. The results from this analysis identifies similarities and differences in the immune responses, particularly soluble cytokines and chemokines, in tissue compartments by using cryptococcal meningitis as a representative model.

## MATERIALS AND METHODS

Subjects with first episode of cryptococcal meningitis were originally enrolled in the Cryptococcal Optimal ART Timing (COAT) trial from November 2010 to April 2012 to determine the optimal time to initiate antiretrovirals after cryptococcal meningitis [3]. The COAT trial was conducted at: Mulago Hospital, Kampala, Uganda; Mbarara University of Science and Technology, Mbarara, Uganda; and the former G.F. Jooste Hospital in Cape Town, South Africa. All subjects completed written informed consent and the institutional review board from each organization approved the study protocol. Details regarding this study trial have been previously described (Clinicaltrials.gov Identifier No: NCT01075152) [3].

We collected peripheral blood and CSF samples at cryptococcal meningitis diagnosis, trial randomization at 1-week, and 2-weeks post cryptococcal meningitis diagnosis. The samples were processed and stored at −80 and then shipped to the University of Minnesota for analysis. Samples did not undergo any prior freeze thaws. Serum and CSF were analyzed in a T cell and macrophage targeted multiplex panel for 19 cytokines and chemokines using a Luminex multiplex cytokine panel (Bio-Rad, Hercules, CA) [4], following the manufacturer’s instructions being run at 1:2 dilution taking the average of two replicates. We also measured CD14 and CD163 by ELISA (R&D Systems, Minneapolis, MN). We have previously reported the longitudinal CSF changes by early vs late ART treatment [4].

To assess the correlation between intra-subject matched serum and CSF cytokines we log transformed the raw readout and subsequently used a non-parametric Spearman correlation test. Data at each time point represents all subjects included in the previous study irrespective of trial arm randomization. Changes in cohort size represents fluctuations in specimen availability, most often due to subjects who declined therapeutic lumbar punctures or subjects who unfortunately died. Significance was set at a p-value of 0.01 to adjust for multiple comparisons in a post-hoc exploratory analysis.

## RESULTS

Of 177 ART-naïve subjects with first episode of cryptococcal meningitis enrolled into the COAT trial, matching CSF and serum existed for 160 subjects at baseline, 105 subjects at 1- week, and 115 subjects at 2-weeks. Missing specimens were most often due to lack of CSF collection on the same day as serum collection as subjects could refuse therapeutic lumbar punctures. Clinical and demographic data comparing COAT trial arms early versus late ART initiation have been previously described [4]. Of the160 subjects included in this analysis, 79 subjects (49%) were female. The median age was 35 (interquartile range (IQR) 28.5 to 40) years with a median CD4 count of 25 (IQR 10 to 74) cells/μL determined at 1-week post diagnosis.

We assessed whether there was a correlation between immune biomarkers in CSF versus serum for 21 analytes irrespective of study arm randomization. At time of diagnosis, none of the 21 immune biomarkers had a positive correlation by Spearman’s non-parametric test (Table 1). Two analytes had statistically significant but weak negative correlations; the type 1 T-helper cell (Th_1_) cytokine interferon-gamma (IFN-γ, Rho= −0.214, p=.007) and the type 2 T-helper cell cytokine (Th_2_) interleukin-4 (IL-4, Rho= −0.232, p=.003). While statistically significant, the correlations were relatively weak and inverse for both IFN-γ and IL-4 (Figure 1), meaning as blood levels increased, CSF levels decreased.

**Table 1:** Correlation of Paired CSF and Serum Analytes at Baseline, 1-Week, and 2-Weeks post Cryptococcal Meningitis Diagnosis.

| Biomarker | At CM diagnosis |  |  | 1-week post diagnosis |  |  | 2-weeks post CM diagnosis |  |  |
| --- | --- | --- | --- | --- | --- | --- | --- | --- | --- |
|  | N | Rho | P-value | N | Rho | P-value | N | Rho | P-value |
| *<u>Innate Immune response</u>* |  |  |  |  |  |  |  |  |  |
| IL-6 | 160 | 0.021 | 0.79 | 105 | -0.025 | 0.80 | 115 | -0.228 | 0.01 |
| IL-1β | 160 | -0.042 | 0.60 | 105 | -0.122 | 0.22 | 115 | 0.052 | 0.58 |
| IL-8 | 160 | -0.034 | 0.67 | 105 | 0.141 | 0.15 | 115 | 0.035 | 0.71 |
| G-CSF | 160 | -0.003 | 0.97 | 105 | 0.1 | 0.31 | 115 | 0.087 | 0.36 |
| GM-CSF | 160 | 0.033 | 0.68 | 105 | 0.16 | 0.10 | 115 | 0.25 | 0.007* |
| MCP-1 | 160 | 0.133 | 0.09 | 105 | 0.216 | 0.03 | 115 | -0.117 | 0.21 |
| MIP-1α | 75 | 0.01 | 0.93 | 5 | 0.4 | 0.51 | 93 | -0.07 | 0.51 |
| MIP-1β | 160 | 0.117 | 0.14 | 105 | 0.114 | 0.25 | 115 | -0.057 | 0.55 |
| CD14 | 87 | 0.078 | 0.47 | n | N/A | N/A | 83 | 0.187 | 0.09 |
| CD163 | 87 | -0.249 | 0.02 | n | N/A | N/A | 83 | -0.099 | 0.38 |
| *<u>Adaptive Immune response</u>* |  |  |  |  |  |  |  |  |  |
| IFN-γ | 160 | -0.214 | 0.007* | 105 | 0.089 | 0.37 | 115 | 0.198 | 0.03 |
| TNF-α | 160 | 0.077 | 0.33 | 105 | 0.038 | 0.70 | 115 | 0.078 | 0.41 |
| IL-12 | 160 | 0.09 | 0.26 | 105 | -0.153 | 0.12 | 115 | -0.005 | 0.96 |
| IL-2 | 160 | -0.025 | 0.76 | 105 | -0.041 | 0.68 | 115 | 0.22 | 0.02 |
| IL-7 | 160 | 0.038 | 0.64 | 105 | 0.11 | 0.26 | 115 | -0.004 | 0.96 |
| IL-10 | 160 | 0.021 | 0.80 | 105 | 0.045 | 0.65 | 115 | -0.025 | 0.80 |
| IL-5 | 160 | -0.046 | 0.56 | 105 | 0.099 | 0.32 | 115 | 0.004 | 0.97 |
| IL-4 | 160 | -0.232 | 0.003* | 105 | 0.107 | 0.28 | 115 | -0.064 | 0.49 |
| IL-13 | 160 | 0.056 | 0.48 | 105 | -0.004 | 0.97 | 115 | -0.057 | 0.54 |
| IL-17 | 160 | -0.008 | 0.92 | 105 | 0.128 | 0.19 | 115 | 0.012 | 0.90 |
| *<u>Hemostasis</u>* |  |  |  |  |  |  |  |  |  |
| VEGF | 111 | -0.049 | 0.61 | 62 | 0.062 | 0.63 | 115 | -0.002 | 0.99 |

\*P value of less than 0.01.
Interleukin (IL), Interferon (IFN), Tumor- necrosis factor (TNF), Granulocyte-colony stimulating factor (G-CSF), Granulocyte-macrophage colony stimulating factor (GM-CSF), Monocyte chemoattractant protein-1/CCL2 (MCP-1), Macrophage inflammatory protein (MIP), and Vascular Endothelial Growth Factor (VEGF)

**Figure 1:**
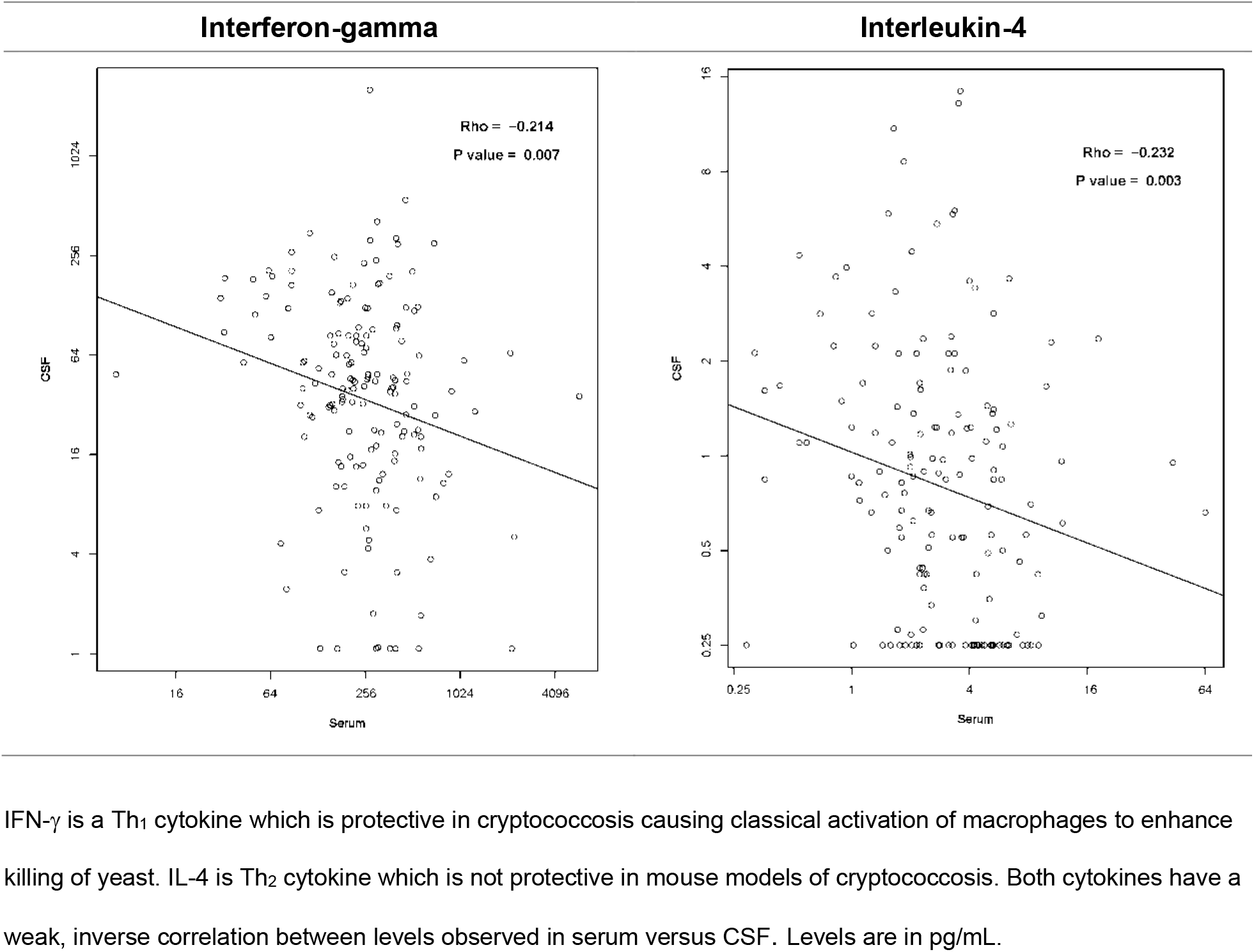
Scatterplot of Interferon-Gamma (IFN-γ) and Interleukin-4 (IL-4) at Cryptococcal Meningitis in CSF Versus Serum.

Of the 19 biomarkers analyzed among 105 paired samples at 1-week post cryptococcal meningitis diagnosis, no cytokines or chemokines were statistically correlated. Of the 21 biomarkers analyzed at 2-weeks post diagnosis among 115 paired samples, a weak positive correlation existed between serum and CSF levels of granulocyte-macrophage colony-stimulating factor (GM-CSF, Rho=0.25, p=.007). Across the three time points as subjects continued standard antifungal therapy, GM-CSF was the only cytokine with a significant correlation between blood and CSF (Rho= 0.147, p=0.003) (Table 2). Additionally, analysis of rate of change over time, through slope calculation across all three time points, demonstrated no significant positive or negative correlation between blood and CSF (Table 2).

**Table 2.**
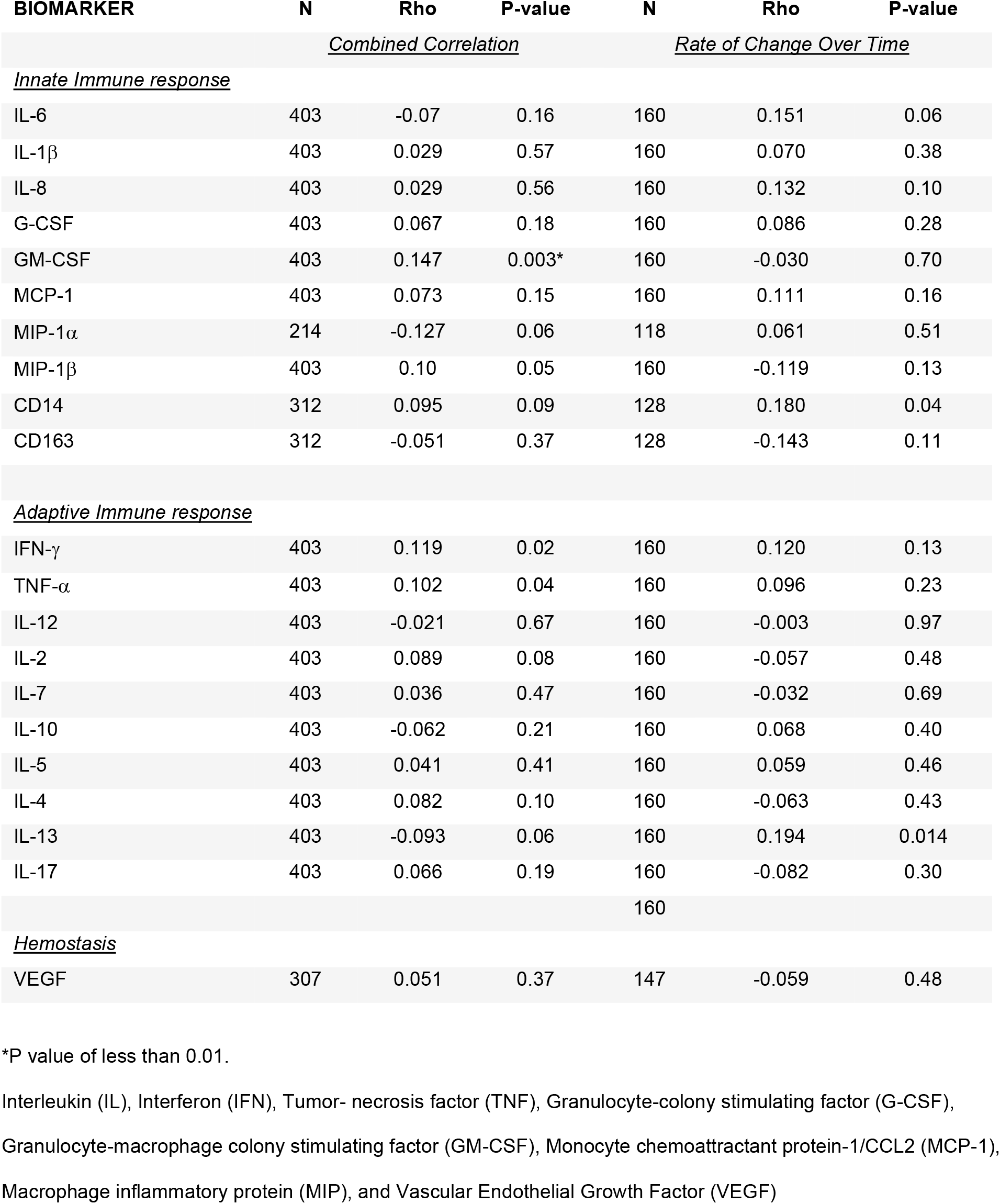
Correlation of CSF and Serum Cytokines Pooled Across All Three Time Points at Baseline, 1-Week, and 2-Weeks post Cryptococcal Meningitis Diagnosis.

## DISCUSSION

We hypothesized that immune biomarkers would have a general positive correlation between serum and CSF. Surprisingly, we found no consistent correlation between soluble immune biomarkers in the peripheral vasculature versus at the site of infection in the CSF. The cytokine environment was strikingly different, even though cryptococcosis is a disseminated infection throughout the vasculature, lungs, and CNS [2, 5, 6]. Additionally, no single analyte had a significant correlation pattern over the course of the first two weeks of standard antifungal therapy. The results from this focused analysis provide evidence that peripheral blood analytes are not necessarily similar to CSF analytes. This builds upon our previous findings which demonstrated that lymphocyte populations and activation levels in CSF are distinct from peripheral blood [7].

There is a large body of published literature comparing human peripheral blood and CSF cytokines and chemokines across various infectious etiologies of meningitis. Comparatively, little literature has been published regarding the correlation within the same individuals’ between human tissue compartments and peripheral blood analytes in infectious etiologies of meningitis [8] or other conditions. Additionally, prior to this study, there was no literature characterizing CSF and blood analyte correlations over time in subjects with cryptococcal meningitis.

Employing cryptococcal meningitis as a model of compartmentalization, we raise a fundamental question of whether the immunology observed in peripheral blood represents what occurs at the end-organ site of infection. This is particularly important as the infectious disease field continues to consider the potential benefits and risks of immunomodulating agent adjuvant therapy, such as administration of proinflammatory cytokines and/or monoclonal antibodies to inhibit anti-inflammatory cytokines or exhaustion receptor engagement. Therefore, it is crucial to truly understand the immune environment at the end-organ site of infection.

The effector immune response at the site of infection appears to be very different from that of cells trafficking in blood. Previous studies on cryptococcal meningitis immune reconstitution inflammatory syndrome documented differential expression of leukocyte phenotype when comparing peripheral blood to the CSF [7]. We doubt these findings are unique to cryptococcosis, for example as demonstrated by differential gene expression when comparing pericardial fluid to peripheral blood in subjects with pericardial tuberculosis [9]. There is also the potential impact of tissue niche on leukocyte activation and cytokine environment.

Research on compartmentalization in infection is limited and thus characterizing the immune response in peripheral blood vs end-organ compartments should be rigorously assessed in other localized infections.

Our study is restricted to immunocompromised persons with cryptococcosis, thus whether the lack of correlation between tissue-specific and peripheral vasculature compartment immune responses is universal, requires future study. Additionally, the repertoire of cytokines and chemokines tested in this analysis include those generated by several T cells subsets, particularly Th1 and Th2 effector T cells, that have been implicated in favorable and detrimental outcomes respectfully in mice with cryptococcal meningitis [10, 11]. The panel also includes several molecules important for macrophage activation and polarization [12] as it has been demonstrated that M1 macrophage polarization enhances *cryptococcus* clearance [13]. Though larger cytokine and chemokine panels are available, this targeted panel includes important analytes that have been consistently studied in cryptococcal meningitis. Therefore, this panel may require modification when analyzing tissue-specific immune responses in other disease states.

It is plausible that participants’ study arm randomization to early or late ART, based on parent clinical trial, could have affected cytokine responses. However, all participants in the parent clinical trial irrespective of study arm randomization, were included in this analysis if CSF and serum compartment matched samples from the same individual were available for analysis. Therefore, it is unlikely that study arm randomization could impact our intra-person correlation analysis. Though 160 subjects were included in the baseline analysis, our data are limited by the absence of 35% and 30% of study participants’ specimens at 1 and 2 weeks, respectively. However, we postulate that more specimens would not have changed the generally poor strength of the correlations. The strength of correlations for all analytes was generally weak with Rho correlation coefficient ranging from −0.23 to +0.25. Thousands of specimens may have been able to generate a statistically significant p-value, but the correlations were weak. Additionally, a more stringent and formal p-value calculation, (for example Bonferroni correction or Benjamini-Hochberg procedure) would not have changed the overall weak correlation.

## CONCLUSION

In conclusion, we demonstrate that cytokines and chemokines released upon leukocyte activation do not correlate between serum and CSF in cryptococcal meningitis. This demonstrates that, though readily available, utilizing peripheral blood as an approximation for the immune status in tissue compartments, such as the CNS, is potentially unreliable.

## Data Availability

All relevant data is included in the manuscript.

## Data Availability

The data used to support the findings in this study are available upon request sent to the corresponding author.

## Acknowledgements

We would like to thank the medical officers, nurses, and laboratory staff who were a part of the COAT Study Team. Additionally, we express our deepest gratitude to all study participants, healthcare professionals, and laboratory staff at all study sites.

## Disclaimer

The findings from this project have not been previously presented at any conferences and is not being considered for publication by another journal at this time.

